# Comparison of Radiomic Feature Aggregation Methods for Patients with Multiple Tumors

**DOI:** 10.1101/2020.11.04.20226159

**Authors:** Enoch Chang, Marina Joel, Hannah Y. Chang, Justin Du, Omaditya Khanna, Antonio Omuro, Veronica Chiang, Sanjay Aneja

**Affiliations:** Department of Therapeutic Radiology, Yale School of Medicine; Yale College; Massachusetts Institute of Technology; Department of Neurosurgery, Thomas Jefferson University; Yale Brain Tumor Program; Department of Neurosurgery, Yale School of Medicine; Center for Outcomes Research and Evaluation, Yale School of Medicine

## Abstract

**Background:** Radiomic feature analysis has been shown to be effective at modeling cancer outcomes. It has not yet been established how to best combine these radiomic features in patients with multifocal disease. As the number of patients with multifocal metastatic cancer continues to rise, there is a need for improving personalized patient-level prognostication to better inform treatment.

**Methods:** We compared six mathematical methods of combining radiomic features of 3596 tumors in 831 patients with multiple brain metastases and evaluated the performance of these aggregation methods using three survival models: a standard Cox proportional hazards model, a Cox proportional hazards model with LASSO regression, and a random survival forest.

**Results:** Across all three survival models, the weighted average of the largest three metastases had the highest concordance index (95% confidence interval) of 0.627 (0.595-0.661) for the Cox proportional hazards model, 0.628 (0.591-0.666) for the Cox proportional hazards model with LASSO regression, and 0.652 (0.565-0.727) for the random survival forest model.

**Conclusions:** Radiomic features can be effectively combined to establish patient-level outcomes in patients with multifocal brain metastases. Future studies are needed to confirm that the volume-weighted average of the largest three tumors is an effective method for combining radiomic features across other imaging modalities and disease sites.

## Introduction

Recent advances in machine learning and diagnostic imaging have increased enthusiasm surrounding the use of quantitative imaging to model clinical outcomes. One quantitative imaging technique that holds particular promise in modeling cancer outcomes is radiomic feature analysis. Radiomic features are quantitative metrics of size, shape, intensity, and texture that are extracted from medical images using high-throughput computational mining techniques.^1,2^ These features represent unique radiographic signatures that have been shown to reveal prognostic insights about underlying gene-expression patterns^3^ and treatment response^4^ that may not be visible to the human eye.

Although radiomic features have shown the ability to risk stratify cancer patients,^5,6^ radiomic analysis has largely been limited to the evaluation of individual tumor volumes. There is no established methodology regarding the best way to combine radiomic features for patients with multifocal sites of disease to establish a patient-level correlate of clinical outcomes. Within oncology, estimating patient outcomes among patients with multiple sites of metastatic disease is of particular interest as it influences multi-disciplinary treatment paradigms.

Prognostication for patients with metastatic disease is an area of increasing need, given as many as 49% of lung cancer patients within the United States are diagnosed with metastatic disease upon initial presentation.^7^ More importantly, nearly 75% of patients with metastatic disease have greater than 5 lesions at diagnosis.^8^ Even among non-metastatic cancer patients, as many as 17% of patients have multi-focal primary tumors.^9^ Brain metastases represent a significant proportion of metastatic cancer patients, and conservatively, at least half of patients diagnosed with metastatic brain disease have multiple brain lesions.^10^ With improved MRI brain imaging, it is likely that the number of patients found with multiple metastases is even higher.^11^

In this study, we attempted to identify optimal techniques to combine radiomic features for patients with multiple brain metastases to model patient-level outcomes. We compared various methods of combining radiomic features of individual tumors in patients with multiple brain metastases and evaluated the performance of different radiomic aggregation methods in estimating survival in patients using different survival models.

## Results

### Patient Demographics

A total of 831 patients with 3596 brain metastases were included. Patient demographics are presented in **Table 1**. Median overall survival was 12 months, median age was 63 years, and the most common primary malignancies were NSCLC (41.0%), melanoma (17.3%), breast (13.1%), SCLC (6.6%), renal (5.9%), and GI (5.3%). 543 patients (65.3%) had <5 metastases, 214 (25.8%) had 5-10 metastases, and 74 (8.9%) had 11+ metastases.

**Table 1.** Patient Demographics

| Age |  |
| --- | --- |
| <50 | 121 (14.6%) |
| 50-59 | 199 (23.9%) |
| 60-69 | 282 (33.9%) |
| >70 | 229 (27.6%) |
| Primary Cancer Site |  |
| NSCLC | 341 (41.0%) |
| Melanoma | 144 (17.3%) |
| Breast | 109 (13.1%) |
| SCLC | 55 (6.6%) |
| Renal | 49 (5.9%) |
| GI | 44 (5.3%) |
| Other | 89 (10.7%) |
| **Number of Metastases** |  |
| <5 | 543 (65.3%) |
| 5-10 | 214 (25.8%) |
| 11+ | 74 (8.9%) |
| **Karnofsky Performance Status\*** |  |
| 100 | 216 (29.1%) |
| 90 | 161 (21.7%) |
| 80 | 160 (21.6%) |
| 70 | 102 (13.8%) |
| 60 | 85 (11.5%) |
| 50 | 13 (1.8%) |
| <=40 | 4 (0.5%) |
| **Presence of Extracranial Metastases** |  |
| Yes | 608 (73.2%) |
| No | 223 (26.8%) |
| * If available |  |

### Overall results

**Table 2** presents a comparison of various aggregation methods across several survival models. For the standard Cox proportional hazards model, the weighted average of the largest 3 metastases had the highest C-index (95% confidence interval) of 0.627 (0.595-0.661). The weighted average of the largest 3 metastases also had the highest C-index of all the aggregation methods on the Cox proportional hazards model with LASSO regression with a C-index of 0.628 (0.591-0.666) and on the random survival forest with a C-index of 0.652 (0.565-0.727).

**Table 2.** Comparison of Aggregation Methods

| **<u>Cox Proportional Hazards</u>** |  |
| --- | --- |
| **<u>Model</u>** | **<u>C-Index (95% CI)</u>** |
| Unweighted Average | 0.610 (0.570-0.646) |
| Weighted Average | 0.604 (0.571-0.641) |
| Weighted Average of Largest 3 Metastases | 0.627 (0.595-0.661) |
| Largest + Number of Metastases | 0.612 (0.579-0.649) |
| Largest Metastasis | 0.598 (0.559-0.636) |
| Smallest Metastasis | 0.595 (0.567-0.631) |
| **<u>Cox Proportional Hazards with LASSO Regression</u>** |  |
| **<u>Model</u>** | **<u>C-Index (95% CI)</u>** |
| Unweighted Average | 0.612 (0.585-0.647) |
| Weighted Average | 0.603 (0.573-0.640) |
| Weighted Average of Largest 3 Metastases | 0.628 (0.591-0.666) |
| Largest + Number of Metastases | 0.597 (0.560-0.632) |
| Largest Metastasis | 0.596 (0.562-0.630) |
| Smallest Metastasis | 0.597 (0.557-0.630) |
| **<u>Random Survival Forest</u>** |  |
| **<u>Model</u>** | **<u>C-Index (95% CI)</u>** |
| Unweighted Average | 0.649 (0.548-0.709) |
| Weighted Average | 0.641 (0.567-0.729) |
| Weighted Average of Largest 3 Metastases | 0.652 (0.565-0.727) |
| Largest + Number of Metastases | 0.622 (0.542-0.706) |
| Largest Metastasis | 0.627 (0.544-0.694) |
| Smallest Metastasis | 0.621 (0.529-0.709) |

### Sub-analysis: Number of Metastases

**Table 3** presents model performance of sub-analyses based on number of brain metastases. For patients with <5 metastases, the weighted average of their 3 largest metastases had the highest C-index on the standard Cox proportional hazards model with a C-index of 0.640 (0.600-0.686). For patients with 5-10 metastases, the unweighted average of all their metastases had the highest C-index of 0.697 (0.638-0.762). For patients with 11+ metastases, the model including only data from the largest metastasis as well as the number of metastases performed with the highest C-index of 0.909 (0.803-0.993).

**Table 3.** Sub-Analysis: Number of Metastases

| **<5 Metastases (n = 543)** |  |
| --- | --- |
| **<u>Model</u>** | **C-Index (95% CI)** |
| Unweighted Average | 0.621 (0.583-0.661) |
| Weighted Average | 0.617 (0.570-0.651) |
| Weighted Average of Largest 3 Metastases | 0.640 (0.600-0.686) |
| Largest + Number of Metastases | 0.639 (0.593-0.676) |
| Largest Metastasis | 0.619 (0.580-0.653) |
| Smallest Metastasis | 0.612 (0.566-0.655) |
| **5-10 Metastases (n=214)** |  |
| **<u>Model</u>** | **C-Index (95% CI)** |
| Unweighted Average | 0.697 (0.638-0.762) |
| Weighted Average | 0.682 (0.617-0.743) |
| Weighted Average of Largest 3 Metastases | 0.688 (0.635-0.749) |
| Largest + Number of Metastases | 0.691 (0.634-0.750) |
| Largest Metastasis | 0.688 (0.623-0.740) |
| Smallest Metastasis | *** |
| **11+ Metastases (n=74)** |  |
| **<u>Model</u>** | **C-Index (95% CI)** |
| Unweighted Average | 0.876 (0.776-0.964) |
| Weighted Average | 0.872 (0.771-0.965) |
| Weighted Average of Largest 3 Metastases | 0.880 (0.787-0.964) |
| Largest + Number of Metastases | 0.909 (0.803-0.993) |
| Largest Metastasis | 0.894 (0.765-0.974) |
| Smallest Metastasis | *** |
| *** Collinear |  |

### Sub-analysis: Volume of Largest Metastasis

**Table 4** describes model performance of sub-analyses based on volume of the largest metastasis. Across all sub-groups, the weighted average of the largest 3 metastases had the highest C-indices of 0.701 (0.652-0.748), 0.687 (0.622-0.728), 0.679 (0.623-0.733) for volumes <0.200 cc, 0.200-0.700 cc, and >0.700+ cc, respectively.

**Table 4.**
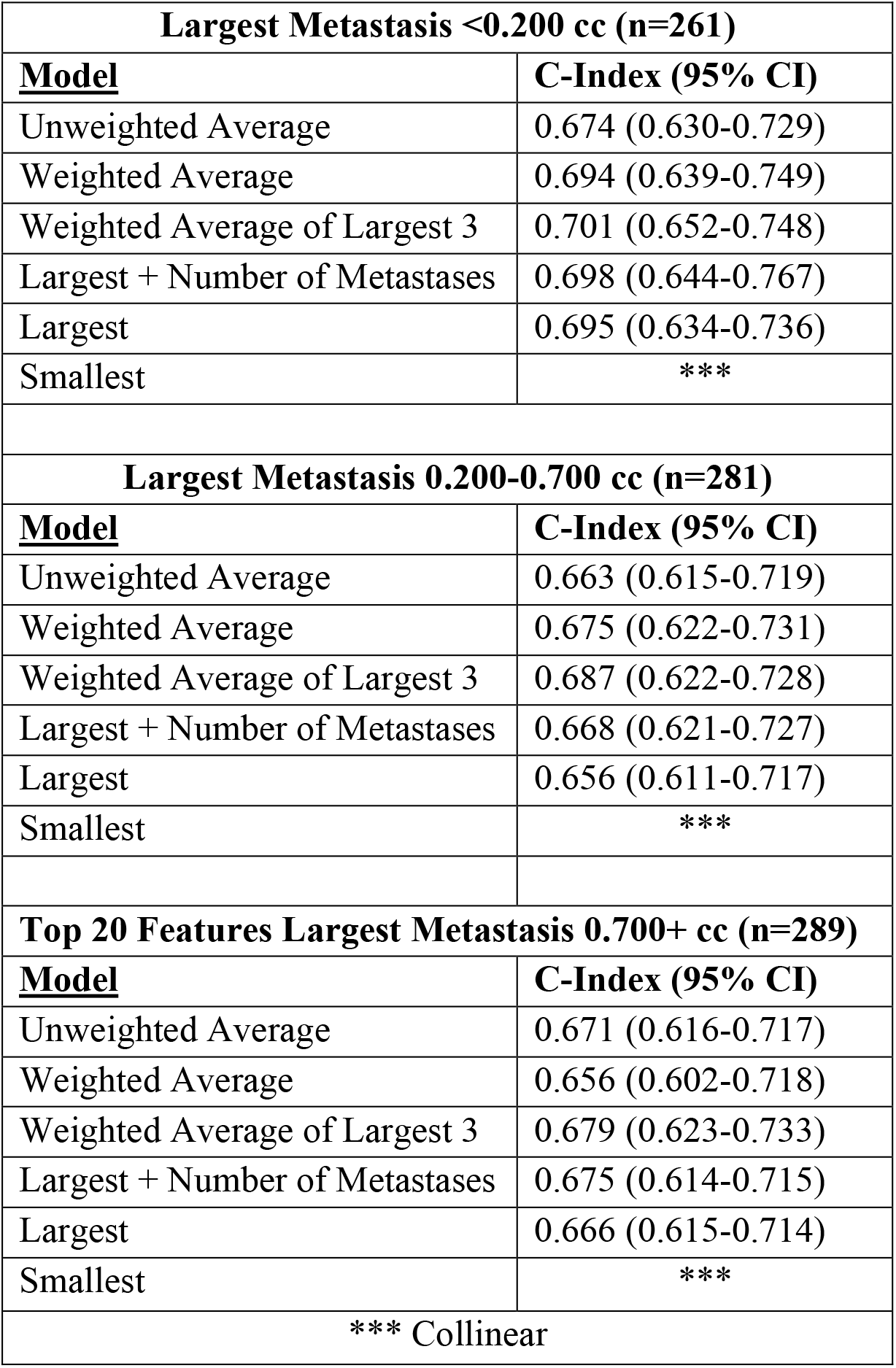
Sub-Analysis: Volume of Largest Metastasis

| **Largest Metastasis <0.200 cc (n=261)** |  |
| --- | --- |
| **<u>Model</u>** | **C-Index (95% CI)** |
| Unweighted Average | 0.674 (0.630-0.729) |
| Weighted Average | 0.694 (0.639-0.749) |
| Weighted Average of Largest 3 | 0.701 (0.652-0.748) |
| Largest + Number of Metastases | 0.698 (0.644-0.767) |
| Largest | 0.695 (0.634-0.736) |
| Smallest | *** |
| **Largest Metastasis 0.200-0.700 cc (n=281)** |  |
| **<u>Model</u>** | **C-Index (95% CI)** |
| Unweighted Average | 0.663 (0.615-0.719) |
| Weighted Average | 0.675 (0.622-0.731) |
| Weighted Average of Largest 3 | 0.687 (0.622-0.728) |
| Largest + Number of Metastases | 0.668 (0.621-0.727) |
| Largest | 0.656 (0.611-0.717) |
| Smallest | *** |
| **Top 20 Features Largest Metastasis 0.700+ cc (n=289)** |  |
| **<u>Model</u>** | **C-Index (95% CI)** |
| Unweighted Average | 0.671 (0.616-0.717) |
| Weighted Average | 0.656 (0.602-0.718) |
| Weighted Average of Largest 3 | 0.679 (0.623-0.733) |
| Largest + Number of Metastases | 0.675 (0.614-0.715) |
| Largest | 0.666 (0.615-0.714) |
| Smallest | *** |
| *** Collinear |  |

## Discussion

The emerging field of radiomic feature analysis has shown particular promise in cancer research. However, traditional radiomic feature analysis has had limited utility for patients with metastatic or multifocal disease because there is a paucity of established methods which aggregate radiomic features across multiple tumors to establish patient-level outcome estimates. Given the majority of cancer patients with brain metastases have multifocal disease, they represent an optimal patient population to study this question. We compared different methods of aggregating radiomic features and tested their performance across different survival models. Our study suggests that a volume-weighted average of the radiomic features of the largest three brain metastases is the most effective technique to model survival across various methods of survival analysis. Furthermore, this suggests that in patients with multi-focal disease, the largest tumors may be driving prognosis.

Practical implications of these findings include more efficient computational and clinical resource utilization. As server costs, data storage requirements, and computation time increase with increasing dataset size and complexity, effective models of prognosis may potentially be developed with just the data from the largest three metastases alone. Furthermore, clinicians may save time by only needing to manually segment the largest three metastases.

This study corroborates studies that have found radiomic features to correlate with prognosis among lung,^12^ breast,^13^ and prostate cancer patients.^6^ To address the issue of combining multi-focal data at a patient-level, prior studies have implemented a weighted average of features of all metastases^14^ while others have included all tumors from a specific patient assigned to either a training or validation set to avoid cluster-correlation biases.^15^ While tumor-level data may be useful for certain tasks like primary-site prediction, there is a need for aggregation of patient-level data for overall outcomes like survival or recurrence.

A notable finding of this study is that across all models, it was important that some measure of multi-focality was incorporated, whether aggregating radiomic features from the three largest metastases or including a clinical measure of multi-focality in the number of metastases.

On sub-analysis, across all volume groups of the largest metastasis, the weighted average of the largest three tumors had the highest performance. As the number of metastases increased greater than 5, models incorporating radiomic data from the largest metastasis with the addition of the number of metastases performed better, suggesting that with a higher number of metastases, additional non-imaging data (i.e. clinical data) may improve model performance, as prior studies have demonstrated the promise of clinical data alone in modeling survival.^16^

This study has limitations. First, there is an inherent selection bias since all the patients in this study were limited to those with brain metastases so results may vary across other disease sites. Second, these patients were all treated at one institution with the same MRI protocol. Third, these patients were treated with the same treatment modality.

In conclusion, this study suggests that radiomic features can be effectively combined to establish patient-level outcomes in patients with multifocal disease. Future studies are needed to confirm that the volume-weighted average of the largest three tumors is an effective method for combining radiomic features across other imaging modalities and disease sites.

## Methods

### Patient Data

Research was conducted in accordance with the Declaration of Helsinki guidelines and approved by the Yale University Institutional Review Board. Informed consent was obtained from all participants in this study. We analyzed 831 patients with 3,596 brain metastases treated with primary stereotactic radiosurgery (SRS) at our institution between 2000-2018. Patients with prior resection or prior radiation treatment were excluded. Metastases <5 mm were also excluded. The primary outcome of interest was overall survival following SRS treatment defined as time from SRS treatment date to date of death or last follow-up.

### Image Preprocessing

The image preprocessing workflow is illustrated in **Figure 1**. Individual metastases were segmented on T1-weighted contrast enhanced MRI images and approved by both a radiation oncologist and a neurosurgeon. A bounding box with 1mm of peri-tumoral tissue around the maximum dimension of the tumor contour in the axial plane was created to ensure edge detection of the tumor. Images were resampled to 1 mm pixel spacing, corrected for low frequency intensity non-uniformity present in MRI data with the N4ITK bias field correction algorithm,^17^ and z-score normalized to reduce inter-scan bias.

**Figure 1.**
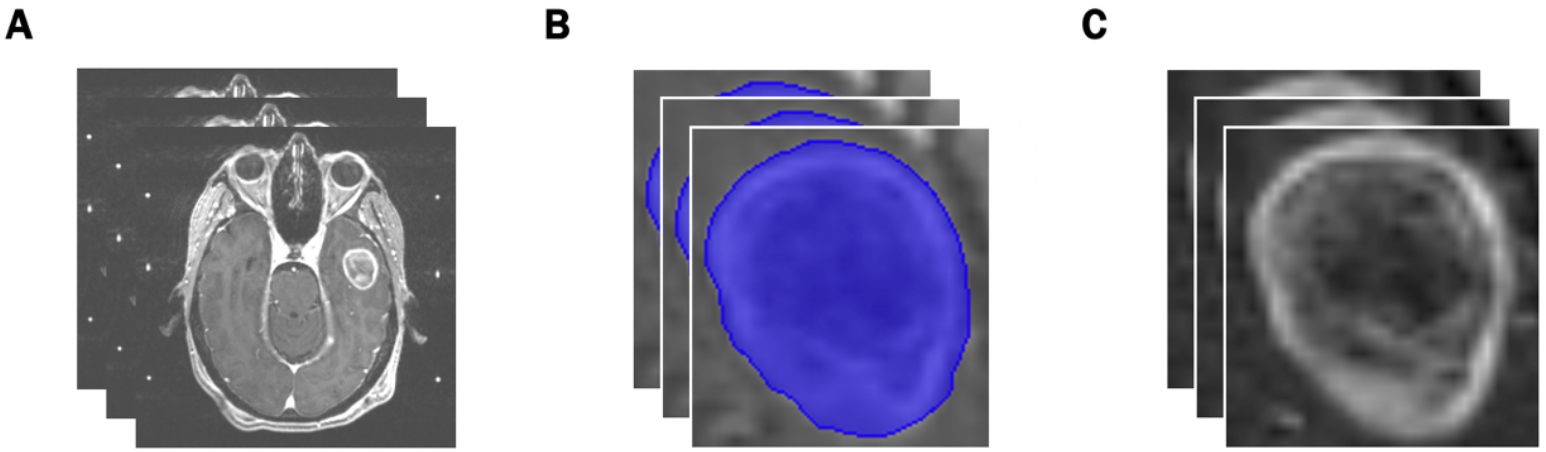
Preprocessing Workflow. **(A)** Input: slices of pre-treatment T1-post contrast brain MRI scans. **(B)** Identification of the region of interest from manual segmentations. **(C)** Output: extracted tumors with pixel resampling, N4ITK bias field correction, and z-score normalization.

### Radiomic Feature Extraction and Aggregation

851 radiomic features^2^ **(Supplement 1)** were extracted from the processed images of each brain metastasis and analyzed as predictors of survival. The following aggregation methods were compared to estimate patient-level risk for patients with multiple metastases: (1) Unweighted Average (UWA): Radiomic size and shape features were summed while all other features were averaged for each patient. (2) Weighted Average (WA): Radiomic size and shape features were summed while all other features were averaged for each patient based on a weighted proportion of total volume of all metastases per patient.^14^ (3) Largest 3 Metastases: Radiomic size and shape features from the largest 3 metastases of each patient were summed while all other features from the largest 3 metastases were averaged based on a weighted proportion of the total volume of the largest 3 metastases.^18^ (4) Largest Metastasis + Number of Metastases: The features from the largest metastasis of each patient were selected, and the total number of metastases for each patient was included as an additional variable.^16^ (5) Largest Metastasis Alone. (6) Smallest Metastasis Alone as a control with the assumption that the smallest tumor would have decreased prognostic value compared to larger tumors.

### Statistical Analysis and Survival Models

Similar to prior radiomic analysis, minimum redundancy maximum relevance (mRMR) was used for dimensionality reduction of radiomic features.^19^ Relevant features were tested with survival models proposed in the literature including a traditional Cox proportional hazards model,^20^ a Cox proportional hazards model with LASSO (least absolute shrinkage and selection operator) regression,^21^ and a random survival forest.^22^ Discriminatory ability of each model was assessed with calculated concordance indices (c-index) using 100 bootstrapped samples of 416 patients (50%). Multiple survival models were assessed to determine if particular aggregation methods were superior for specific survival models.

### Sub-Group Analysis

A sub-group analysis was performed to evaluate the role of the following potential drivers of patient prognosis: the overall number of metastases^16^ and the volume of the largest metastasis of each patient.^18^ The robustness of the various radiomic aggregation techniques was evaluated by comparing the Cox proportional hazards model across these sub-groups: the number of metastases (<5 metastases, 5-10 metastases, 11+ metastases) as well as the volume of the largest metastasis (<0.200 cc, 0.200-0.700 cc, > 0.700+ cc) per patient.

## Data Availability

Data is available upon request.

## Code Availability

Radiomic features were extracted using PyRadiomics (version 2.2.0).^23^ Minimum redundancy maximum relevance feature selection was performed with the mRMRe package using R version 3.6.2.^24^ Image preprocessing and comparison of aggregation were performed with Python version 3.7. Survival analysis was performed with the lifelines and scikit-survival packages.^25^ Our code is available at https://github.com/Aneja-Lab-Yale.

## Acknowledgments

SA: Research funding from the American Cancer Society (ACS-IRG 17-172-57). EC: Research funding from the Radiological Society of North America (RMS1904), Yale Center for Clinical Investigation (NIH CTSA TL1 TR001864), James G. Hirsch Medical Research Fellowship.

## Author Contributions

Conception and Design: EC, SA. Data Collection: EC, MJ, VC. Data Analysis and Interpretation: All Authors. Drafting the Manuscript: EC, SA. Critical Revision: All Authors. Final Approval: All Authors.

## Table and Figure Legend

**Supplemental Table S1**. Radiomic Features

**Supplemental S1.**
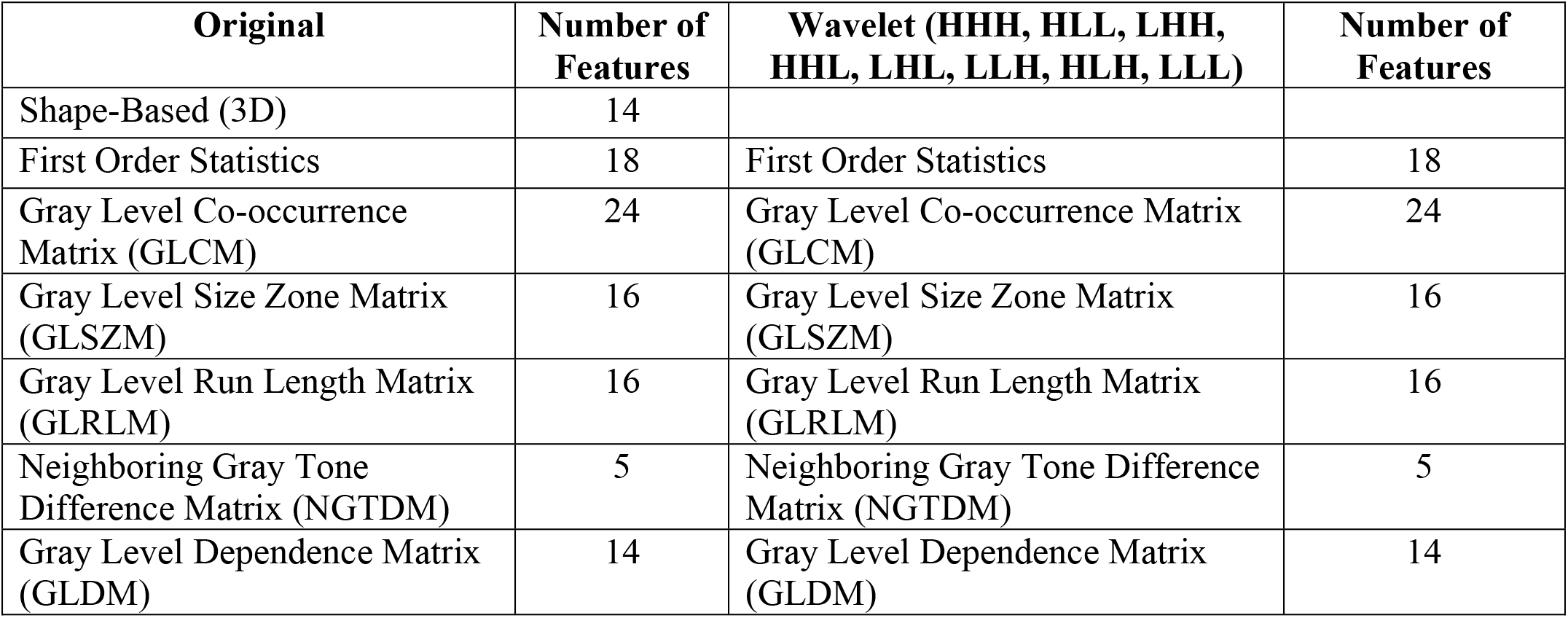
Radiomic Features The following Original feature classes were extracted using the Pyradiomics package.^1^ Features for each of the Wavelet filter classes (HHH, HLL, LHH, HHL, LHL, LLH, HLH, LLL) were also extracted for a total of 851 extracted radiomic features.

## References

1 Kumar, V. et al. Radiomics: the process and the challenges. Magn Reson Imaging 30, 1234–1248, doi:10.1016/j.mri.2012.06.010 (2012).

2 Lambin, P. et al. Radiomics: the bridge between medical imaging and personalized medicine. Nat Rev Clin Oncol 14, 749–762, doi:10.1038/nrclinonc.2017.141 (2017).

3 Aerts, H. J. et al. Decoding tumour phenotype by noninvasive imaging using a quantitative radiomics approach. Nat Commun 5, 4006, doi:10.1038/ncomms5006 (2014).

4 Dercle, L. et al. Identification of Non-Small Cell Lung Cancer Sensitive to Systemic Cancer Therapies Using Radiomics. Clin Cancer Res, doi:10.1158/1078-0432.CCR-19-2942 (2020).

5 Kickingereder, P. et al. Radiomic subtyping improves disease stratification beyond key molecular, clinical, and standard imaging characteristics in patients with glioblastoma. Neuro Oncol 20, 848–857, doi:10.1093/neuonc/nox188 (2018).

6 Osman, S. O. S. et al. Computed Tomography-based Radiomics for Risk Stratification in Prostate Cancer. Int J Radiat Oncol Biol Phys 105, 448–456, doi:10.1016/j.ijrobp.2019.06.2504 (2019).

7 National Cancer Institute Cancer Trends Progress Report: Stage Distribution, <https://progressreport.cancer.gov/diagnosis/stage> (2020).

8 Parikh, R. B. et al. Definitive primary therapy in patients presenting with oligometastatic non-small cell lung cancer. Int J Radiat Oncol Biol Phys 89, 880–887, doi:10.1016/j.ijrobp.2014.04.007 (2014).

9 Vogt, A. et al. Multiple primary tumours: challenges and approaches, a review. ESMO Open 2, e000172, doi:10.1136/esmoopen-2017-000172 (2017).

10 Expert Panel on Radiation Oncology-Brain, M. et al. American College of Radiology appropriateness criteria on multiple brain metastases. Int J Radiat Oncol Biol Phys 75, 961–965, doi:10.1016/j.ijrobp.2009.07.1720 (2009).

11 Nagai, A. et al. Increases in the number of brain metastases detected at frame-fixed, thin-slice MRI for gamma knife surgery planning. Neuro Oncol 12, 1187–1192, doi:10.1093/neuonc/noq084 (2010).

12 Hosny, A. et al. Deep learning for lung cancer prognostication: A retrospective multi-cohort radiomics study. PLoS Med 15, e1002711, doi:10.1371/journal.pmed.1002711 (2018).

13 Park, H. et al. Radiomics Signature on Magnetic Resonance Imaging: Association with Disease-Free Survival in Patients with Invasive Breast Cancer. Clin Cancer Res 24, 4705–4714, doi:10.1158/1078-0432.CCR-17-3783 (2018).

14 Dercle, L. et al. Radiomics Response Signature for Identification of Metastatic Colorectal Cancer Sensitive to Therapies Targeting EGFR Pathway. J Natl Cancer Inst, doi:10.1093/jnci/djaa017 (2020).

15 Kniep, H. C. et al. Radiomics of Brain MRI: Utility in Prediction of Metastatic Tumor Type. Radiology 290, 479–487, doi:10.1148/radiol.2018180946 (2019).

16 Sperduto, P. W. et al. Summary report on the graded prognostic assessment: an accurate and facile diagnosis-specific tool to estimate survival for patients with brain metastases. J Clin Oncol 30, 419–425, doi:10.1200/JCO.2011.38.0527 (2012).

17 Tustison, N. J. et al. N4ITK: improved N3 bias correction. IEEE Trans Med Imaging 29, 1310–1320, doi:10.1109/TMI.2010.2046908 (2010).

18 Weltman, E. et al. Radiosurgery for brain metastases: a score index for predicting prognosis. Int J Radiat Oncol Biol Phys 46, 1155–1161, doi:10.1016/s0360-3016(99)00549-0 (2000).

19 Ding, C. & Peng, H. Minimum redundancy feature selection from microarray gene expression data. J Bioinform Comput Biol 3, 185–205, doi:10.1142/s0219720005001004 (2005).

20 Cox, D. R. Regression Models and Life-Tables. Journal of the Royal Statistical Society. Series B (Methodological) 34, 187–220 (1972).

21 Tibshirani, R. The lasso method for variable selection in the Cox model. Stat Med 16, 385–395, doi:10.1002/(sici)1097-0258(19970228)16:4<385::aid-sim380>3.0.co;2-3 (1997).

22 Ishwaran, H., Kogalur, U. B., Blackstone, E. H. & Lauer, M. S. Random survival forests. Ann. Appl. Stat. 2, 841–860, doi:10.1214/08-AOAS169 (2008).

23 van Griethuysen, J. J. M. et al. Computational Radiomics System to Decode the Radiographic Phenotype. Cancer Res 77, e104–e107, doi:10.1158/0008-5472.CAN-17-0339 (2017).

24 De Jay, N. et al. mRMRe: an R package for parallelized mRMR ensemble feature selection. Bioinformatics 29, 2365–2368, doi:10.1093/bioinformatics/btt383 (2013).

25 Pölsterl, S. “scikit-survival: A Library for Time-to-Event Analysis Built on Top of scikit-learn” Journal of Machine Learning Research 21, 1–6 (2020).

## References

1 van Griethuysen, J. J. M. et al. Computational Radiomics System to Decode the Radiographic Phenotype. Cancer Res 77, e104–e107, doi:10.1158/0008-5472.CAN-17-0339 (2017).

